# Leveraging Generative AI to Accelerate Biocuration of Medical Actions for Rare Disease

**DOI:** 10.1101/2024.08.22.24310814

**Authors:** Enock Niyonkuru, J Harry Caufield, Leigh C Carmody, Michael A Gargano, Sabrina Toro, Patricia L. Whetzel, Hannah Blau, Mauricio Soto Gomez, Elena Casiraghi, Leonardo Chimirri, Justin T Reese, Giorgio Valentini, Melissa A Haendel, Christopher J Mungall, Peter N. Robinson

## Abstract

Structured representations of clinical data can support computational analysis of individuals and cohorts, and ontologies representing disease entities and phenotypic abnormalities are now commonly used for translational research. The Medical Action Ontology (MAxO) provides a computational representation of treatments and other actions taken for the clinical management of patients. Currently, manual biocuration is used to assign MAxO terms to rare diseases, enabling clinical management of rare diseases to be described computationally for use in clinical decision support and mechanism discovery. However, it is challenging to scale manual curation to comprehensively capture information about medical actions for the more than 10,000 rare diseases.

We present AutoMAxO, a semi-automated workflow that leverages Large Language Models (LLMs) to streamline MAxO biocuration for rare diseases. AutoMAxO first uses LLMs to retrieve candidate curations from abstracts of relevant publications. Next, the candidate curations are matched to ontology terms from MAxO, Human Phenotype Ontology (HPO), and MONDO disease ontology via a combination of LLMs and post-processing techniques. Finally, the matched terms are presented in a structured form to a human curator for approval. We used this approach to process 4,918 unique medical abstracts and identified annotations for 21 rare genetic diseases, we extracted 18,631 candidate disease-treatment curations, 538 of which were confirmed and transferred to the MAxO annotation dataset.

The results of this project underscore the potential of generative AI to accelerate precision medicine by enabling a robust and comprehensive curation of the primary literature to represent information about diseases and procedures in a structured fashion. Although we focused on MAxO in this project, similar approaches could be taken for other biomedical curation tasks.

## Introduction

Rare diseases present significant challenges in healthcare due to their low prevalence and high complexity. While individual rare diseases have low prevalence, collectively they affect approximately one in ten Americans and nearly 300 million people globally (1). Most rare conditions are difficult to diagnose, with many patients waiting years for accurate diagnosis. Despite over 10,000 rare diseases having been identified, fewer than 5% have FDA-approved treatments (2,3). Clinicians and researchers depend on various resources such as GeneReviews, primary literature, clinical trial databases, and other published medical literature to identify potential treatments. These resources, however, present a disjointed landscape that can be burdensome to navigate effectively. Biomedical ontologies are structured frameworks that categorize medical concepts to enable sophisticated searching and algorithmic analysis, and can thereby support clinicians and researchers in finding relevant information.

Large language models (LLMs) are advanced AI models trained on extensive text data to interpret and generate human language. These models use deep learning techniques to produce contextually relevant and coherent text, demonstrating versatility in translation, summarization, and question-answering tasks. LLMs have been applied to numerous medical domains (4). Biocuration, the process of collecting, organizing, and annotating biological data, ensures the accuracy and usefulness of information for research and clinical applications. In the realm of biological research, accurate data curation is crucial for meaningful scientific advancements. Biocurators manually review literature and databases to extract data about genes, proteins, diseases, and phenotypes, organizing it into structured ontologies (5). However, biocuration requires significant human effort, highlighting the need for automated solutions to enhance efficiency in biocuration (6).

To reduce the workload of manual curators, various efforts have developed methods for completing information extraction tasks with LLMs. Agarwal et al. found that OpenAI’s GPT-3 model could accurately complete specific extraction tasks, such as abbreviation expansion and medication attribute extraction, from clinical text with no specific training. They also found that prompts directing the LLM to yield a specific output structure improved performance in resolving results (7). Since that time, information extraction researchers have assembled both new benchmark sets (e.g., MultiMedQA (4), BioLLMBench (8)) and domain-adapted models (e.g., Med-PaLM (4), Med-MLLM (9), BioMistral (10)) to extract an increasingly broad range of data elements from medical text. SPIRES (Structured Prompt Interrogation and Recursive Extraction of Semantics) (11) is a knowledge extraction method that leverages both LLMs and ontologies. SPIRES uses knowledge schemas defined using LinkML (12) to extract entities and relationships from text. Since each schema encapsulates specific domain concepts, relationships, and properties, it can be used to craft more effective prompts for LLMs, therefore obtaining reliable annotations in the form of textual elements. A key aspect of SPIRES’ effectiveness is its ability to ground these textual elements as concepts derived from ontologies (i.e., to identify ontology terms that match the concepts identified by the LLM), including Open Biological and Biomedical Ontologies (OBO) Foundry ontologies (13).

The Medical Action Ontology (MAxO) provides a computational representation of medical diagnostics, preventions, procedures, interventions, and therapies. MAxO follows OBO standards, with terms having unique identifiers, names/labels definitions, in addition to computational logical definitions, and synonyms that can be used for NLP applications. A medical action is broadly regarded as any medical procedure, intervention, therapy, and or measurement undertaken for clinical management. The structure of MAxO is composed of six upper-level terms, viz., diagnostic procedure, preventative therapy, therapeutic procedure, medical action avoidance, palliative care, and complementary, and alternative medical therapy (14). Currently, MAxO includes 1,902 medical action terms, curated through manual and semi-automated methods. MAxO is compatible with other ontologies within the OBO Foundry (13), enhancing the ability to model diseases and phenotypic features comprehensively. It provides a computational representation of treatments and actions for clinical patient management and is integrated with the Mondo Disease Ontology (Mondo) and the Human Phenotype Ontology (HPO), broadening the scope of computational disease modeling of rare diseases (14).

Like many ontologies, in OBO, MAxO is used for the annotation of knowledge curated from the literature. The MAxO annotation model is designed to systematically describe medical actions and their relationships to diseases and phenotypic features. It enables a structured and interoperable way to represent clinical interventions and management strategies within biomedical research and healthcare by capturing relationships between disease, phenotypes, and medical action. The core MAxO annotation model relates medical actions to phenotypes in the context of a disease For example:

> Medical Action: *copper chelator agent therapy [MAXO:0001224]*
>
> Relationship: PREVENTS
>
> Phenotype: *Cirrhosis [HP:0001394]*
>
> Disease: *Wilson disease Anemia [MONDO:0010200]*

Identifiers are used for all concepts to avoid ambiguity. MAxO annotations are disease-specific. In this example, copper chelator agent therapy is indicated to prevent Cirrhosis in the context of Wilson disease Anemia. However, *copper chelator agent therapy [MAXO:0001224]* would not be indicated in other diseases characterized by *Cirrhosis [HP:0001394]*, such as *Hepatitis C [MONDO:0005231], Primary Biliary Cholangitis [MONDO:0005388]* or *Alcoholic Liver Disease [MONDO:0043693]*

Additional examples of MAxO annotations are provided in Table 1. Not shown here, but the model also allows for the specification of full provenance and evidence, including the publication from which the annotation was derived from. The current source of truth for MAxO annotations is a tabular file maintained on GitHub (https://github.com/monarch-initiative/maxo-annotations).

**Table 1.** Summary of the counts of novel annotations derived with automaxo. 538 novel annotations were obtained for a total of 21 diseases that involved a total of 152 unique HPO terms and 114 unique MAxO terms. One example annotation is shown for each disease.

| MONDO | Annotat<br>ions | MAxO (n) | HPO (n) | Example MAxO | Example<br>relation | Example HPO |
| --- | --- | --- | --- | --- | --- | --- |
| Achondroplasia<br>(MONDO:0007037) | 2 | 1 | 1 | human growth hormone<br>replacement therapy<br>(MAXO:0000780) | prevents | Short stature (HP:0004322) |
| alkaptonuria<br>(MONDO:0008753) | 13 | 7 | 6 | pharmacotherapy<br>(MAXO:0000058) | treats | Elevated urinary<br>homogentisic acid<br>(HP:0033704) |
| Apert syndrome<br>(MONDO:0007041) | 24 | 11 | 15 | airway management<br>(MAXO:0000500) | treats | Apert syndrome<br>(MONDO:0007041) |
| Brugada syndrome<br>(MONDO:0015263) | 10 | 3 | 6 | ablation therapy<br>(MAXO:0000452) | prevents | Ventricular arrhythmia<br>(HP:0004308) |
| Camurati-Engelmann<br>disease<br>(MONDO:0007542) | 10 | 7 | 5 | corticosteroid agent therapy<br>(MAXO:0000640) | treats | Bone pain (HP:0002653) |
| Canavan disease<br>(MONDO:0010079) | 8 | 4 | 4 | gene therapy<br>(MAXO:0001001) | treats | Canavan disease<br>(MONDO:0010079) |
| celiac disease<br>(MONDO:0005130) | 67 | 1 | 8 | dietary gluten intake<br>avoidance<br>(MAXO:0010000) | prevents | celiac disease<br>(MONDO:0005130) |
| Chediak-Higashi<br>syndrome<br>(MONDO:0008963) | 17 | 3 | 2 | allogeneic hematopoietic<br>stem cell transplantation<br>(MAXO:0001479) | treats | Chediak-Higashi syndrome<br>(MONDO:0008963) |
| citrullinemia, type II,<br>adult-onset<br>(MONDO:0011326) | 14 | 5 | 3 | carbohydrate-restricted diet<br>intake (MAXO:0000771) | treats | Hepatic encephalopathy<br>(HP:0002480) |
| Donnai-Barrow<br>syndrome<br>(MONDO:0009104) | 8 | 7 | 8 | surgical procedure<br>(MAXO:0000004) | treats | Congenital diaphragmatic<br>hernia (HP:0000776) |
| Huntington disease<br>(MONDO:0007739) | 46 | 22 | 16 | palliative care<br>(MAXO:0000021) | treats | Huntington disease<br>(MONDO:0007739) |
| hypochondroplasia<br>(MONDO:0007793) | 4 | 3 | 3 | human growth hormone<br>replacement therapy<br>(MAXO:0000780) | treats | Short stature (HP:0004322) |
| Lesch-Nyhan syndrome<br>(MONDO:0010298) | 21 | 9 | 9 | umbilical cord blood<br>transplantation<br>(MAXO:0010033) | treats | Lesch-Nyhan syndrome<br>(MONDO:0010298) |
| Loeys-Dietz syndrome<br>(MONDO:0018954) | 20 | 11 | 15 | gastrostomy<br>(MAXO:0001346) | treats | Failure to thrive<br>(HP:0001508) |
| lymphatic malformation<br>1 (MONDO:0007919) | 1 | 1 | 1 | physical therapy<br>(MAXO:0000011) | treats | Lymphedema<br>(HP:0001004) |
| Marfan syndrome<br>(MONDO:0007947) | 44 | 17 | 18 | angiotensin receptor<br>blocker therapy<br>(MAXO:0000653) | prevents | Aortic root aneurysm<br>(HP:0002616) |
| Noonan syndrome<br>(MONDO:0018997) | 31 | 17 | 22 | behavioral dietary<br>intervention<br>(MAXO:0000884) | treats | Gastroparesis<br>(HP:0002578) |
| propionic acidemia<br>(MONDO:0011628) | 22 | 8 | 4 | pharmacotherapy<br>(MAXO:0000058) | treats | Acute hyperammonemia<br>(HP:0008281) |
| sickle cell anemia<br>(MONDO:0011382) | 117 | 31 | 21 | gene therapy<br>(MAXO:0001001) | treats | sickle cell anemia<br>(MONDO:0011382) |
| Stickler syndrome<br>(MONDO:0019354) | 11 | 6 | 5 | vitrectomy<br>(MAXO:0001085) | treats | Rhegmatogenous retinal<br>detachment (HP:0012230) |
| Wilson disease<br>(MONDO:0010200) | 48 | 9 | 16 | liver transplantation<br>(MAXO:0001175) | treats | Hepatic failure<br>(HP:0001399) |

The MAxO annotation model is currently being mapped to the Biolink Model, to allow for inclusion into Knowledge Graphs (KG), such as the NCATS Translator KG, Monarch KG, and KG-Hub, and to allow graph machine learning methods such as link prediction to apply to these data using software such as GRAPE.

Currently, MAxO annotation is a manual process, involving an expert curator selecting and carefully reading relevant papers and manually entering annotations via a specialized tool. We present AutoMAxO, a semi-automated workflow that leverages LLMs and SPIRES to assist with the annotation and update of the MAxO ontology for rare diseases.

## Methodology

### Automated retrieval of relevant text

To automatically mine the relevant literature from PubMed, AutoMAxO either uses a list of PubMed identifiers (PMID) representing published articles selected by the users or exploits a selection of MeSH codes representing diseases of interest and then employs the NCBI E-Utilities API (15) to gather PubMed IDs of peer-reviewed articles focused on these diseases. By using specific search criteria, the user may tailor the search to, for example, retrieve articles related to potential therapies, use disease keywords and MeSH tree mappings to filter the results or provide a maximum number of papers to be retrieved, or prioritize the retrieval of publications that are within time-ranges or being the most relevant (highly cited). The user specifies the number of articles to retrieve. To avoid duplication, AutoMAxO first checks the existing directory to see if any articles have already been retrieved. Then, it retrieves the exact number of additional articles specified by the user. Once the maximum number of articles available has been reached, the number of articles saved is less than what the user desired, which means the articles relevant to the particular disease and the keywords mentioned above have reached the maximum.

After retrieving relevant PubMed IDs from E-Utilities that were not extracted before, AutoMAxO uses PubTator 3.0 API to retrieve titles and abstracts that correspond to the PubMed IDs (16). AutoMAxO saves the texts in a comma-separated values (CSV) file, to be used as input for the next step of LLM-driven parsing.

### Structured prompt interrogation and recursive extraction of semantics (SPIRES)

For extracting structured information from the text, we utilized OntoGPT Version 0.3.9, a Python package we developed for parsing text with large language models (LLMs), using instruction prompts and ontology-based grounding. Within OntoGPT, we used GPT-4 model version ‘gpt-4-0125-preview’ to extract annotations related to medical actions, relationships with diseases, and phenotypes. OntoGPT implements SPIRES (11), which incorporates a schema and ontology-driven extraction of annotations from text. In this process, using our MAxO schema (https://github.com/monarch-initiative/automaxo/blob/main/src/automaxo/maxo_template.yaml), OntoGPT generates candidate annotations consisting of five elements: subjects (medical actions), predicates (relationships), objects (phenotypes), qualifiers (diseases), and subject extensions (chemical entities). The title and abstract of each article were used as separate inputs for OntoGPT and the extracted data were saved in YAML format. This YAML contains objects conforming to the AutoMAxO schema, using term identifiers from relevant ontologies. Where OntoGPT cannot ground terms in ontologies, it makes placeholder terms (i.e., it reports words or phrases that might correspond to a term from the corresponding target ontology; the curator can replace the placeholder with the term, if one exists, or create a new term request for a concept that is not currently represented in the ontology).

Each file is further post-processed to identify close lexical matches between extracted terms and those in MAxO. Through the post-processing, we also use the annotation feature of the Ontology-Access Kit (OAK) to match the subset of terms that were not already grounded with the existing ontologies using lexical matching of characters and words. For example, OntoGPT may identify *allogeneic bone marrow transplantation* as a potential MAxO term, but this term has no exact match within MAxO. OAK locates two potential MAxO terms related to the extracted annotation: *MAxO:0010030 (bone marrow transplantation)* and *MAxO:0000068 (transplantation)*. These are broader than the concepts needed to fully capture the annotation, but they are valid for use.

After further grounding and identifying potential ontology terms, AutoMAxO combines and groups all extracted annotations, ranking them by their frequency of occurrence. For each annotation, AutoMAxO records literature evidence, including PubMed IDs and relevant text excerpts from which the annotations were extracted. All results are saved in a JSON file that can be used by curation tools. By default, AutoMAxO stores result in a folder called “data”, and 20 such folders are included in the GitHub repository as examples. A subfolder is created for each analyzed disease. For instance, one of the subfolders is called alkaptonuria. Each disease folder contains several files created by AutoMAxO. The file called “final_automaxo_results.json” in each disease folder is the one that should be used for curation tools.

### Automaxoviewer

We created a JavaFX graphical user interface (GUI)-based tool called Automaxoviewer that presents the results of AutoMAxO (from the final_automaxo_results.json file) in tabular form and provides autocomplete and various other functions for a curator to validate and if needed correct or extend the results of AutoMAxO.

For instance, if AutoMAxO is not able to ground a term and provide an exact ontology term, but instead returns a lexical variant (e.g., “liver transpl.” instead of *liver transplantation [MAxO:0001175])*, the curator can use the autocomplete functionality to assign the MAxO term.

After careful vetting, the annotation can be directly added to the main MAxO annotation database on GitHub. The Automaxoviewer code is freely available on GitHub under a GNU General Public License version 3 open-source license at https://github.com/monarch-initiative/automaxoviewer,

## Results

AutoMAxO is an approach towards streamlining the curation of treatments and other medical actions in the medical literature about rare diseases. AutoMAxO first collects candidate abstracts from PubMed, then it leverages OntoGPT to extract structured information from each abstract with GPT-4. This information is further processed to identify ontology term identifiers for the concepts (“grounding”) and to output a JSON file with the results. Finally, candidate annotations are presented to a domain expert for vetting (Figure 2).

**Figure 1.**
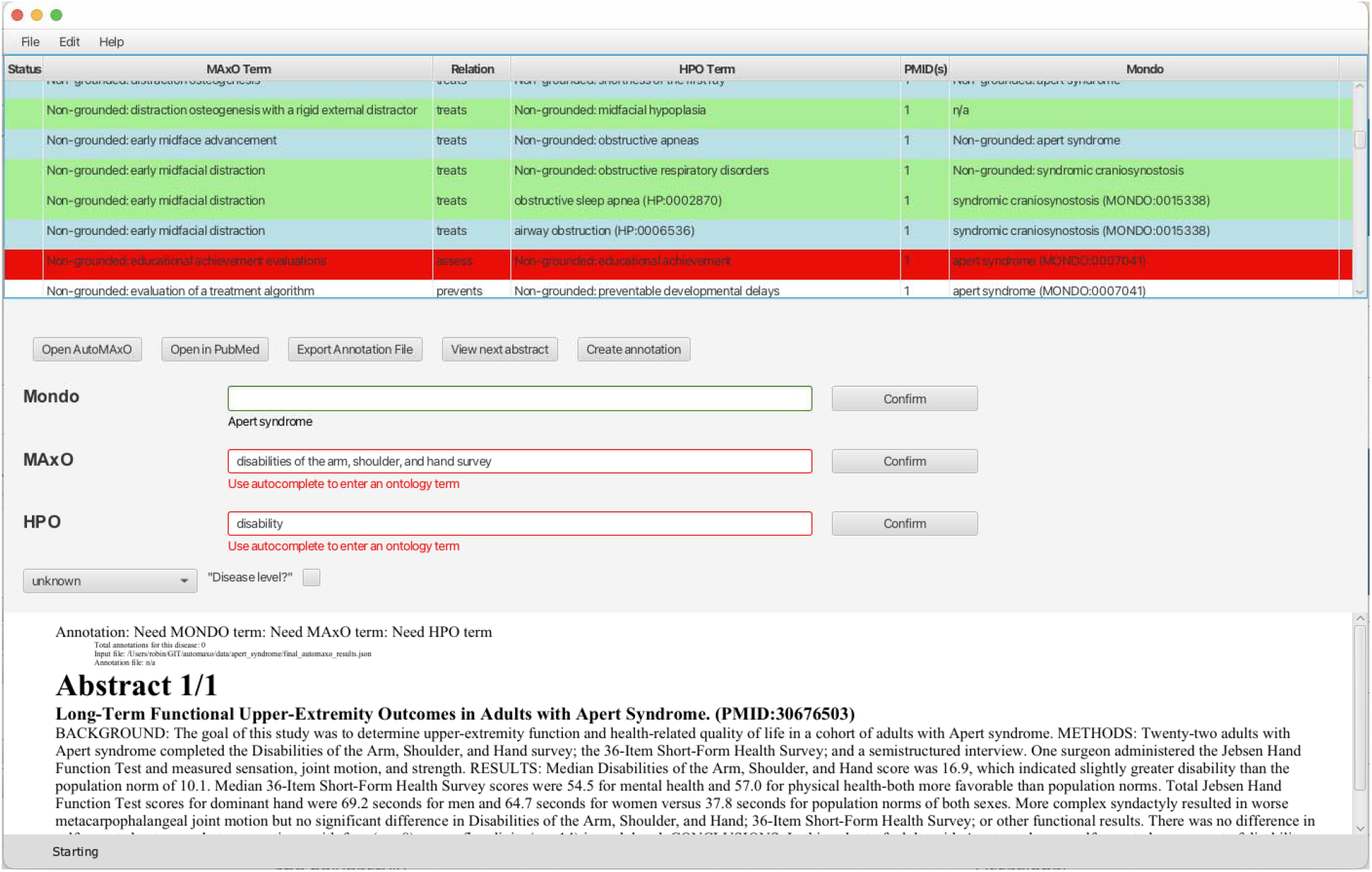
Screenshot of the Automaxoviewer application used to screen the results from AutoMAxO. The last author (PNR; board-certified pediatrician with Habilitation in human genetics) reviewed candidate annotations for relevance and medical correctness.

**Figure 2:**
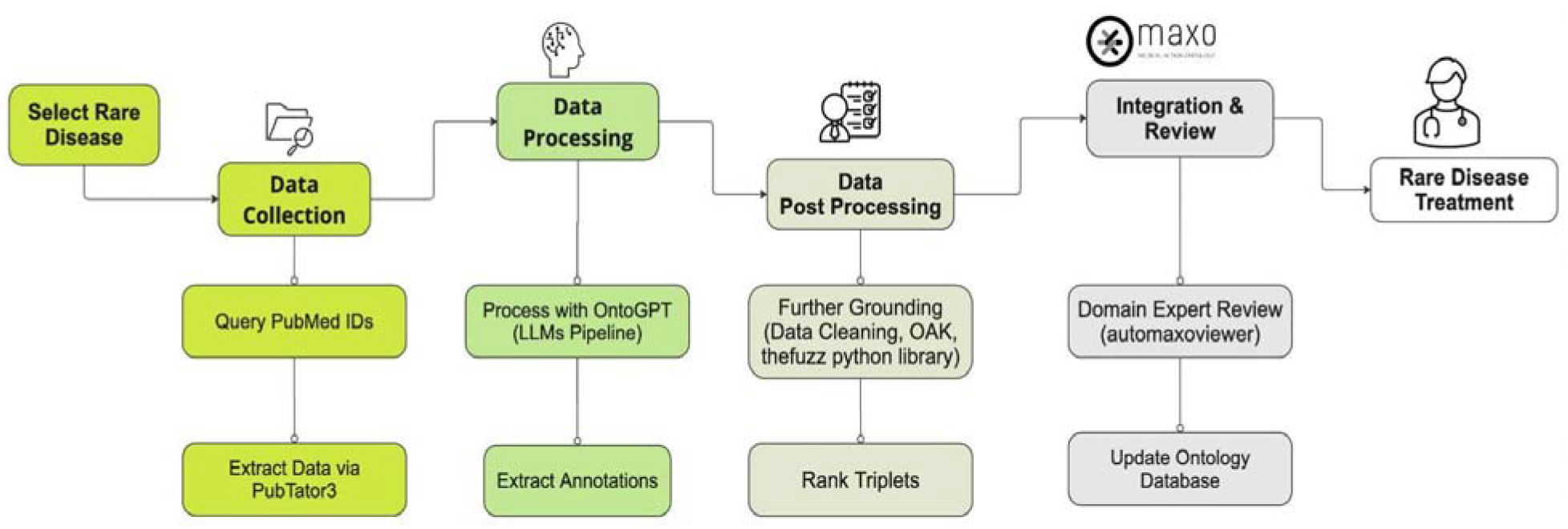
Workflow Diagram for AutoMAxO Project: This diagram illustrates the step-by-step process of the AutoMAxO project, from selecting rare diseases and collecting data to processing, validating, and integrating data into the MAxO database for enhancing rare disease treatment.

AutoMAxO was evaluated on 21 rare genetic diseases that were randomly selected, and 4,918 abstracts were retrieved and passed to OntoGPT for further processing. AutoMAxO proposed 18,631 unique candidate annotations, which were summarized and provided to MAxO curators for review to update the MAxO database. The candidates were reviewed using Automaxoviewer, and annotations were confirmed if they described case reports, cohort reports, clinical studies, or reviews about medical actions applied to individuals with the disease in question. Articles that described other aspects of the diseases, such as genetic studies, molecular mechanisms, or animal models, were excluded, as were retrieved candidates that did not specifically discuss the target disease. This enabled a total of 538 novel annotations to be confirmed for a total of 21 rare diseases. A summary of the new annotations is provided in Table 1, and a full list is available as Supplemental File 1. The annotations have additionally been added to the MAxO annotation GitHub repository for download.

## Discussion

In this work, we have presented an LLM-based approach to accelerate curation using three current bio-ontologies: MAxO, HPO, and MONDO. AutoMAxO automates several curation tasks that would otherwise be time-consuming and labor-intensive for manual curators, such as identifying abstracts to curate and suggesting relevant ontology terms. AutoMAxO successfully extracted a total of 18,631 unique annotations for 21 rare genetic diseases.

We have currently implemented AutoMAxO to emphasize recall over precision. Many of the returned abstracts are related to the disease but do not represent reports of medical actions. For instance, some returned abstracts described research with model organisms, genetic diagnostics, or other topics (Figure 3). Using AutoMAxO Viewer, it is straightforward to scan the title and abstract text to skip such entries. We are currently testing machine-learning approaches to filter the abstracts to improve precision. In some cases, abstracts could not be annotated because a MAxO term was lacking. In this case, AutoMAxO Viewer automatically generates text for a new term request and opens the MAxO issues page in GitHub to streamline the process.

**Figure 3:**
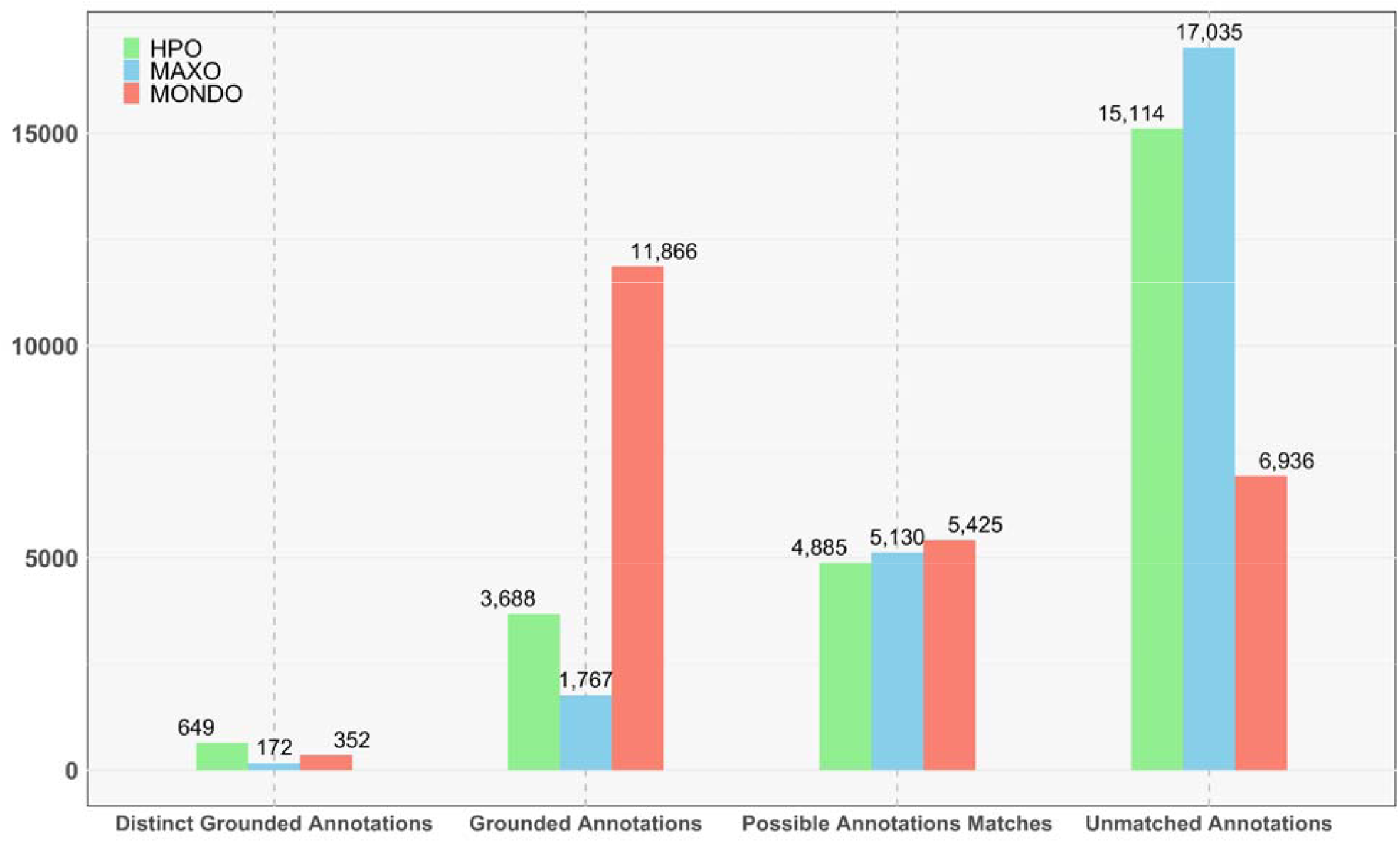
This figure summarizes the annotations extracted from 4,918 PubMed entries by the AutoMAxO tool. It presents the counts of Distinct Annotations (all terms identified at least once), Grounded Annotations (all entities extracted from the text and successfully mapped to a MAxO, HPO, or MONDO identifier), Possible Annotation Matches (all entities extracted from the text and with a predicted mapping to an ontology identifier based on lexical similarity), and Unmatched Annotations (all medical actions identified by the LLM but unable to be mapped to an ontology term) for MAxO, HPO, or MONDO.

The June 2023 release of the existing MAxO annotation database included 1,757 MAxO terms,(14) 18,490 HPO terms, and 411 MONDO terms. AutoMAxO significantly enhanced the speed and efficiency of the curation process. With just 21 sample diseases analyzed in our study, we curated 538 new annotations with AutoMAxO and merged them with our existing MAxO annotation files. Although it is challenging to objectively compare the time required for each annotation, our previous efforts resulted in 438 annotations over a five-year period for the MAxO resource, primarily through a manual process performed by curators, which was time-consuming. In contrast, with AutoMAxO, the annotation of each disease was completed in approximately one to two hours. This remarkable improvement underscores the potential of leveraging LLMs to streamline and expedite the curation process.

AutoMAxO integrates LLMs and advanced search technologies to automate the annotation and updating of medical ontologies. Existing models such as BioBERT(17) support NLP and curation methods for specific tasks like Named Entity Recognition (NER) and relationship extraction, but they generally need extensive training data to extract medical actions. We are not aware of any other tools currently available tool that specifically retrieves concepts from multiple ontologies (here, MAxO, Mondo, and HPO) that are related by multiple rules (in MAxO, treatments are related to phenotypic features and diseases by the relations treats, prevents, investigates, contraindicated, and lack of observed response).

Additionally, with minimal configuration, AutoMAxO can extract annotations from custom texts, including full PubMed Central texts, websites, or other text collections. AutoMAxO’s ability to access the latest research literature to extract annotations using ontologies ensures that medical data remains up-to-date, supporting more effective and targeted patient care. A similar approach could be applied to other curation tasks in biomedicine or other fields by adapting the SPIRES template file that specifies the schema with ontology terms and relations.

## Conclusion

In this study, we describe and evaluate AutoMAxO, a workflow for annotating published reports of treatments and other medical actions for rare diseases. AutoMAxO streamlines several time-consuming steps in the process of curation and has enabled us to double the number of annotations for disease-specific MAxO annotations.

## Data Availability

All data produced are available online at https://github.com/monarch-initiative/automaxo.

https://github.com/monarch-initiative/automaxo

## Code availability

The source code for AutoMAxO is available on GitHub at: https://github.com/monarch-initiative/automaxo.

Documentation for AutoMAxO is available here:

https://monarch-initiative.github.io/automaxo/

The source code for the AutoMAxO Viewer is available on GitHub at: https://github.com/monarch-initiative/automaxoviewer

